# SARS-CoV-2 seroprevalence in the urban population of Qatar: An analysis of antibody testing on a sample of 112,941 individuals

**DOI:** 10.1101/2021.01.05.21249247

**Authors:** Peter V. Coyle, Hiam Chemaitelly, Mohamed Ali Ben Hadj Kacem, Naema Hassan Abdulla Al Molawi, Reham Awni El Kahlout, Imtiaz Gilliani, Nourah Younes, Zaina Al Kanaani, Abdullatif Al Khal, Einas Al Kuwari, Adeel A. Butt, Andrew Jeremijenko, Anvar Hassan Kaleeckal, Ali Nizar Latif, Riyazuddin Mohammad Shaik, Hanan F. Abdul Rahim, Gheyath K. Nasrallah, Hadi M. Yassine, Mohamed G. Al Kuwari, Hamad Eid Al Romaihi, Mohamed H. Al-Thani, Roberto Bertollini, Laith J. Abu-Raddad

## Abstract

**Background:** Qatar has experienced a large SARS-CoV-2 epidemic. Our first objective was to assess the proportion of the urban population that has been infected with SARS-CoV-2, by measuring the prevalence of detectable antibodies. Our second objective was to identify predictors for infection and for having higher antibody titers.

**Methods:** Residual blood specimens from individuals receiving routine and other clinical care between May 12-September 9, 2020 were tested for anti-SARS-CoV-2 antibodies. Associations with seropositivity and higher antibody titers were identified through regression analyses. Probability weights were applied in deriving the epidemiological measures.

**Results:** We tested 112,941 individuals (∼10% of Qatar’s urban population), of whom 51.6% were men and 66.0% were 20-49 years of age. Seropositivity was 13.3% (95% CI: 13.1-13.6%) and was significantly associated with sex, age, nationality, clinical-care type, and testing date. The proportion with higher antibody titers varied by age, nationality, clinical-care type, and testing date. There was a strong correlation between higher antibody titers and seroprevalence in each nationality, with a Pearson correlation coefficient of 0.85 (95% CI: 0.47-0.96), suggesting that higher antibody titers may indicate repeated exposure to the virus. The percentage of antibody-positive persons with prior PCR-confirmed diagnosis was 47.1% (95% CI: 46.1-48.2%), severity rate was 3.9% (95% CI: 3.7-4.2%), criticality rate was 1.3% (95% CI: 1.1-1.4%), and fatality rate was 0.3% (95% CI: 0.2-0.3%).

**Conclusions:** Fewer than two in every 10 individuals in Qatar’s urban population had detectable antibodies against SARS-CoV-2 between May 12-September 9, 2020, suggesting that this population is still far from the herd immunity threshold and at risk from a subsequent epidemic wave.

## Introduction

With the breakthrough development of highly efficacious vaccines against the severe acute respiratory syndrome coronavirus 2 (SARS-CoV-2) [1-3], determining the population’s cumulative infection exposure and current immunity level is critical to inform national vaccine roll-out strategies.

Qatar, located in the Arabian peninsula, with a multinational population of 2.8 million people, nearly all living in the capital city, Doha, had a significant first wave of COVID-19 that peaked in late May 2020 [4,5]. As of December 23, 2020, >60,000 infections per million population had been laboratory-confirmed [6,7]. Qatar has a unique socio-demographic structure, in which single-unit and family households including children, adults and/or older adults, account for only 40% of the total population, with adults in this “urban population” often being part of the professional or service workforce [4,8,9]. The remaining 60% of the population consists of craft and manual workers (CMWs) [4,8,9]—mostly single, young men working in development projects [9] and typically living in large, shared accommodations [10].

Infection transmission in Qatar was first documented among CMWs on March 6, 2020 [11], who were subsequently most affected by this epidemic [12]. A recently completed nationwide, population-based survey assessing “ever” infection among the CMW population found that six out of every ten persons had detectable antibodies against SARS-CoV-2, suggesting that this population is at or near herd immunity [13]. In the present study, the first objective was to assess the level of infection exposure among the rest of the population of Qatar, that of the “urban population” of this country. The second objective was to identify predictors for infection and for having higher antibody titers.

## Methods

### Data sources

A cross-sectional serological study was conducted from May 12 to September 9, 2020 to assess the level of and associations with antibody positivity in the urban population of Qatar. The sample included residual blood specimens collected from individuals receiving routine and other clinical care at Hamad Medical Corporation (HMC), a main provider of healthcare to the urban population of this country and the nationally designated provider for Coronavirus Disease 2019 (COVID-19) healthcare needs.

Antibody data generated during the study were subsequently linked to the national centralized SARS-CoV-2 polymerase chain reaction (PCR) testing and hospitalization database, which includes records for all PCR testing and COVID-19 hospitalizations in Qatar since the start of the epidemic [14]. The database further includes the severity classification of hospitalized cases, based on individual chart reviews completed by trained medical personnel using the World Health Organization (WHO) criteria [15]. The study was approved by the HMC and Weill Cornell Medicine-Qatar Institutional Review Boards. The study was conducted following the ethics review boards guidelines and regulations.

### Laboratory methods

Roche Elecsys^®^ Anti-SARS-CoV-2 (Roche, Switzerland), an electrochemiluminescence immunoassay, was used for antibody detection in serological samples. Result interpretation followed manufacturer instructions: reactive for optical density (a proxy for antibody titer) cutoff index ≥1.0 and non-reactive for cutoff index <1.0 [16].

Current infection was assessed using PCR testing of aliquots of Universal Transport Medium (UTM) used for nasopharyngeal and oropharyngeal swab collection (Huachenyang Technology, China). Aliquots were extracted on the QIAsymphony platform (QIAGEN, USA) and tested with real-time reverse-transcription PCR (RT-qPCR) using the TaqPath™ COVID-19 Combo Kit (Thermo Fisher Scientific, USA) on an ABI 7500 FAST (Thermo Fisher, USA). Samples were extracted using a custom protocol [17] on a Hamilton Microlab STAR (Hamilton, USA) and tested using the AccuPower SARS-CoV-2 Real-Time RT-PCR Kit (Bioneer, Korea) on an ABI 7500 FAST, or loaded directly into a Roche cobas® 6800 system and assayed with the cobas® SARS-CoV-2 Test (Roche, Switzerland). All laboratory testing was conducted at HMC Central Laboratory following standardized protocols.

### Statistical analysis

Frequency distributions were used to describe sample characteristics and optical density among antibody-positive persons. Probability weights were applied to generate estimates representing the wider urban population. Weights were developed using population distributions by sex, age group, and nationality in the Primary Health Care Corporation (PHCC) database [18]. This database essentially covers the urban population of Qatar and includes 1,468,837 registered users, distributed across Qatar’s 27 PHCC centers [18].

Associations with anti-SARS-CoV-2 positivity, as well as with higher antibody titers (defined as optical density higher than the median value) were investigated using chi-square tests and univariable logistic regression. Covariates with p-values ≤0.2 in univariable regression analysis were included in the multivariable model. Covariates with p-values ≤0.05 in the multivariable analysis were regarded as strong evidence for an association with the outcome. Odds ratios (ORs), adjusted ORs (AORs), 95% confidence intervals (CIs), and p-values were reported.

The antibody database was linked to the SARS-CoV-2 PCR testing and hospitalization database to enable estimation of other epidemiologic metrics. The latter included the proportion of antibody-positive persons who had a diagnosis of SARS-CoV-2 confirmed by PCR prior to the antibody test. Numbers of infections that were classified as severe, critical, or fatal, according to WHO criteria [15], among all antibody-positive persons, were used to estimate severity, criticality, and fatality rates.

## Results

In all, 112,941 individuals were tested, representing ∼10% of the urban population of Qatar [4] (Table 1). Of these, 51.6% were men. Two-thirds (66%) of tested persons were 20-49 years of age. Qatari (25.8%) and Indian nationals (16.5%) were most heavily represented in the sample, reflecting their representation in the urban population [5,8,19]. Specimens were collected in the course of routine clinical care during home care visits (34.2%), outpatient visits (28.5%), inpatient hospital stays (21.0%), and emergency department visits (16.4%). Overall, the sample mirrored the urban population demographics [5,8,9].

**Table 1.**
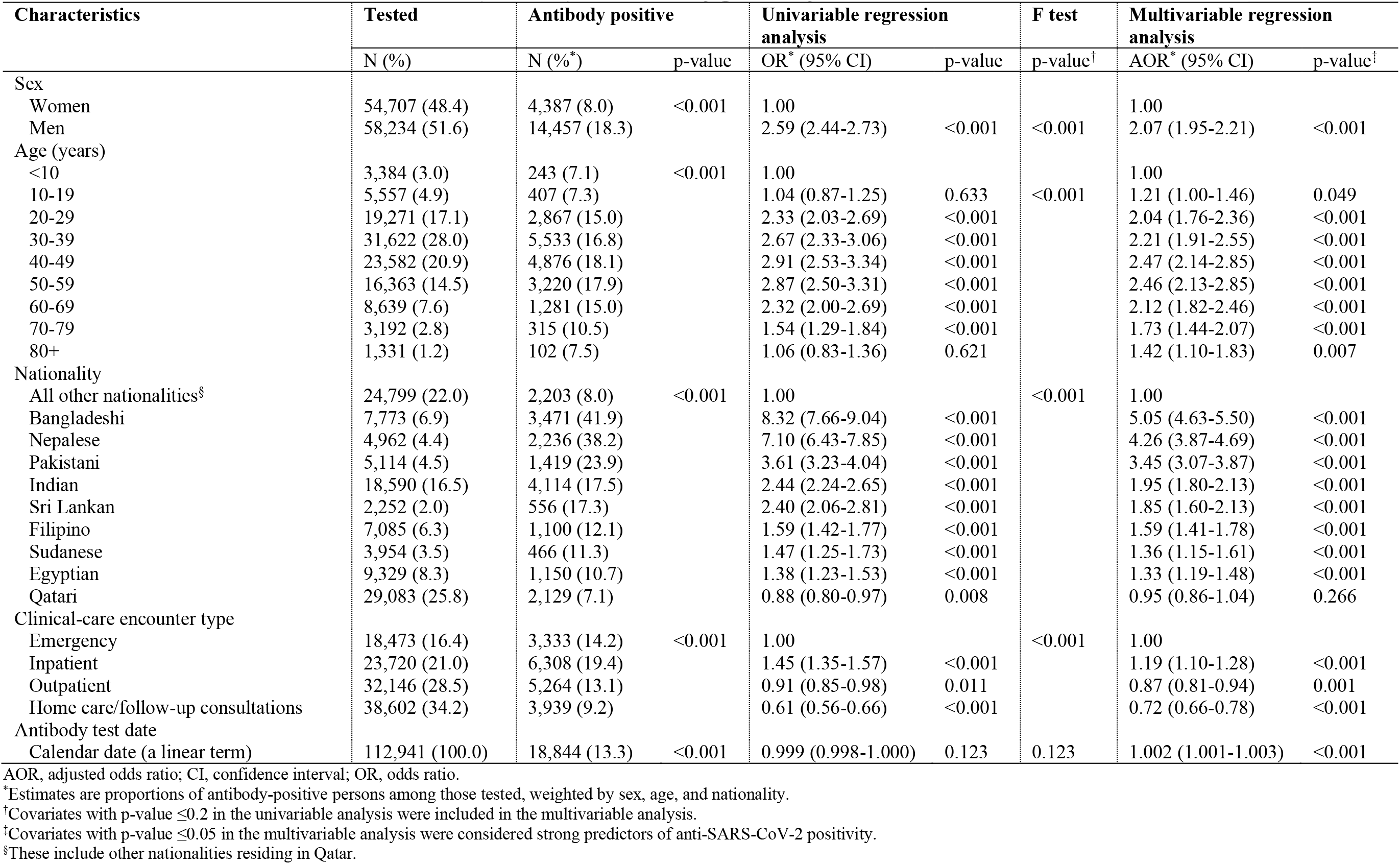
Characteristics of tested individuals (112,941) and antibody positivity.

A total of 18,844 individuals had detectable SARS-CoV-2 antibodies—a weighted antibody positivity of 13.3% (95% CI: 13.1-13.6%). Seropositivity was independently associated with sex, age, nationality, clinical-care type, and calendar date of the antibody test in the multivariable regression analysis (Table 1). Men had two-fold higher odds of being seropositive (AOR of 2.07; 95% CI: 1.95-2.21) than women. Similarly, the AOR was two-fold higher for adults 20-79 years of age than for children <10 years of age. Seropositivity varied by nationality. Compared to other nationalities, AOR was 5.05 (95% CI: 4.63-5.50) for Bangladeshis, 4.26 (95% CI: 3.87-4.69) for Nepalese, 3.45 (95% CI: 3.07-3.87) for Pakistanis, 1.95 (95% CI: 1.80-2.13) for Indians, 1.85 (95% CI: 1.60-2.13) for Sri Lankans, 1.59 (95% CI: 1.41-1.78) for Filipinos, 1.36 (95% CI: 1.15-1.61) for Sudanese, 1.33 (95% CI: 1.19-1.48) for Egyptians, and 0.95 (95% CI: 0.86-1.04) for Qataris. Compared to emergency department attendees, AOR was 0.87 (95% CI: 0.81-0.94) for outpatients and 0.72 (95% CI: 0.66-0.78) for patients with home-care visits or follow-up consultations, and 1.19 (95% CI: 1.10-1.28) for inpatients. There was evidence of increasing seropositivity over time (Table 1 and Table S1 of Supplementary Information (SI)), but at a slow rate. The AOR (per day) was 1.002 (95% CI: 1.001-1.003; Table 1).

Figure 1 illustrates the distribution of antibody titers (optical density values) among the 18,844 antibody-positive persons. Optical density values ranged from 1.0 to 150.0 with a median of 27.0. Having higher antibody titers than the median was not associated with sex, but in the multivariable regression analysis they were independently associated with age, nationality, clinical-care type, and the calendar date of the antibody test (Table 2). Compared to those aged 20-29 years, the AOR was higher in children <10 years and adults aged 40-79 years. There were significant differences by nationality. AOR was 1.68 (95% CI: 1.45-1.94) for Bangladeshis, 1.54 (95% CI: 1.32-1.80) for Nepalese, 1.30 (95% CI: 1.05-1.61) for Filipinos, 1.22 (95% CI: 1.05-1.43) for Indians, 1.19 (95% CI: 0.98-1.44) for Pakistanis, 1.12 (95% CI: 0.87-1.44) for Sri Lankans, 1.04 (95% CI: 0.77-1.41) for Sudanese, 0.82 (95% CI: 0.67-1.01) for Egyptians, and 0.78 (95% CI: 0.65-0.94) for Qataris. Compared to emergency department attendees, inpatients had an AOR for higher antibody positivity of 0.38 (95% CI: 0.34-0.43), while no difference was found for outpatients or for patients with home-care visits or follow-up consultations. Having higher antibody titers increased with time (Table 2 and Table S2 of SI), with an AOR (per day) of 1.011 (95% CI: 1.010-1.013; Table 2).

**Table 2.**
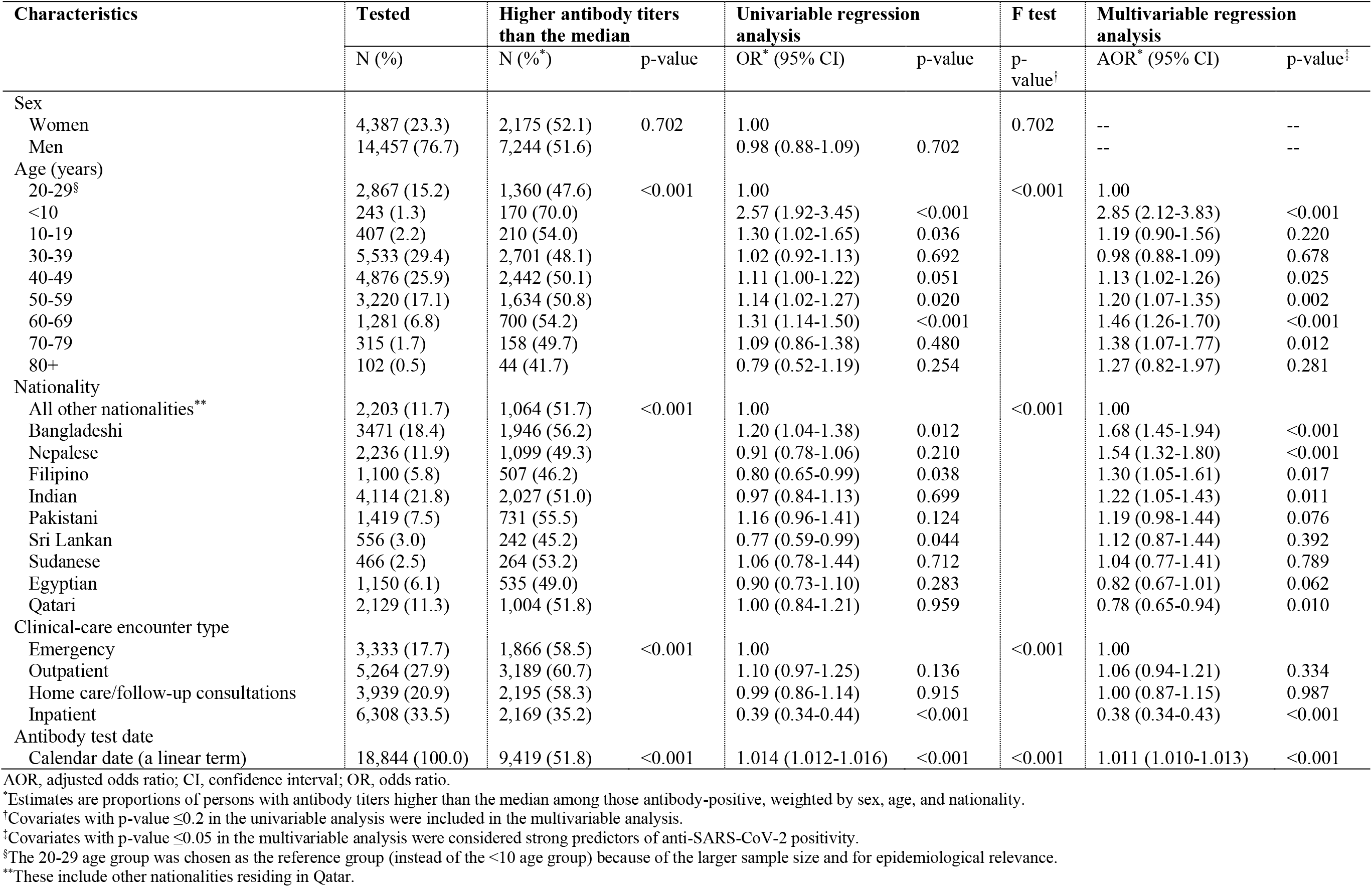
Associations of antibody titers (optical densities) higher than the median value (≥27.0) among the 18,844 antibody-positive individuals.

**Figure 1.**
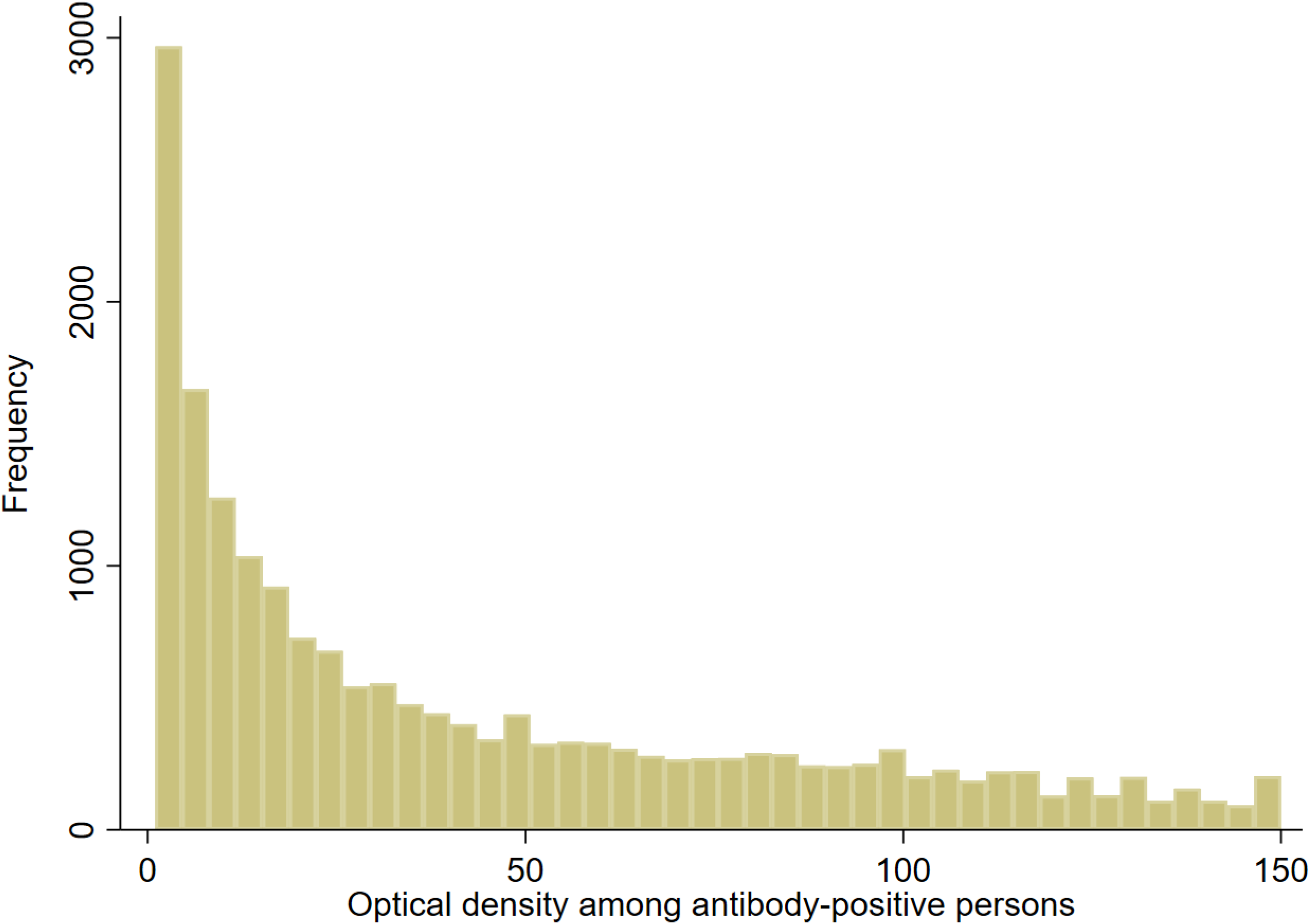
Distribution of antibody titers (optical density values) among the 18,844 antibody-positive individuals identified in this study.

There was a strong correlation between the AOR for higher antibody titers in each nationality and the corresponding SARS-CoV-2 seroprevalence of that nationality (Figure 2). The Pearson correlation coefficient was 0.85 (95% CI: 0.47-0.96), possibly indicating that higher antibody titers correlate with repeated exposures to this coronavirus.

**Figure 2.**
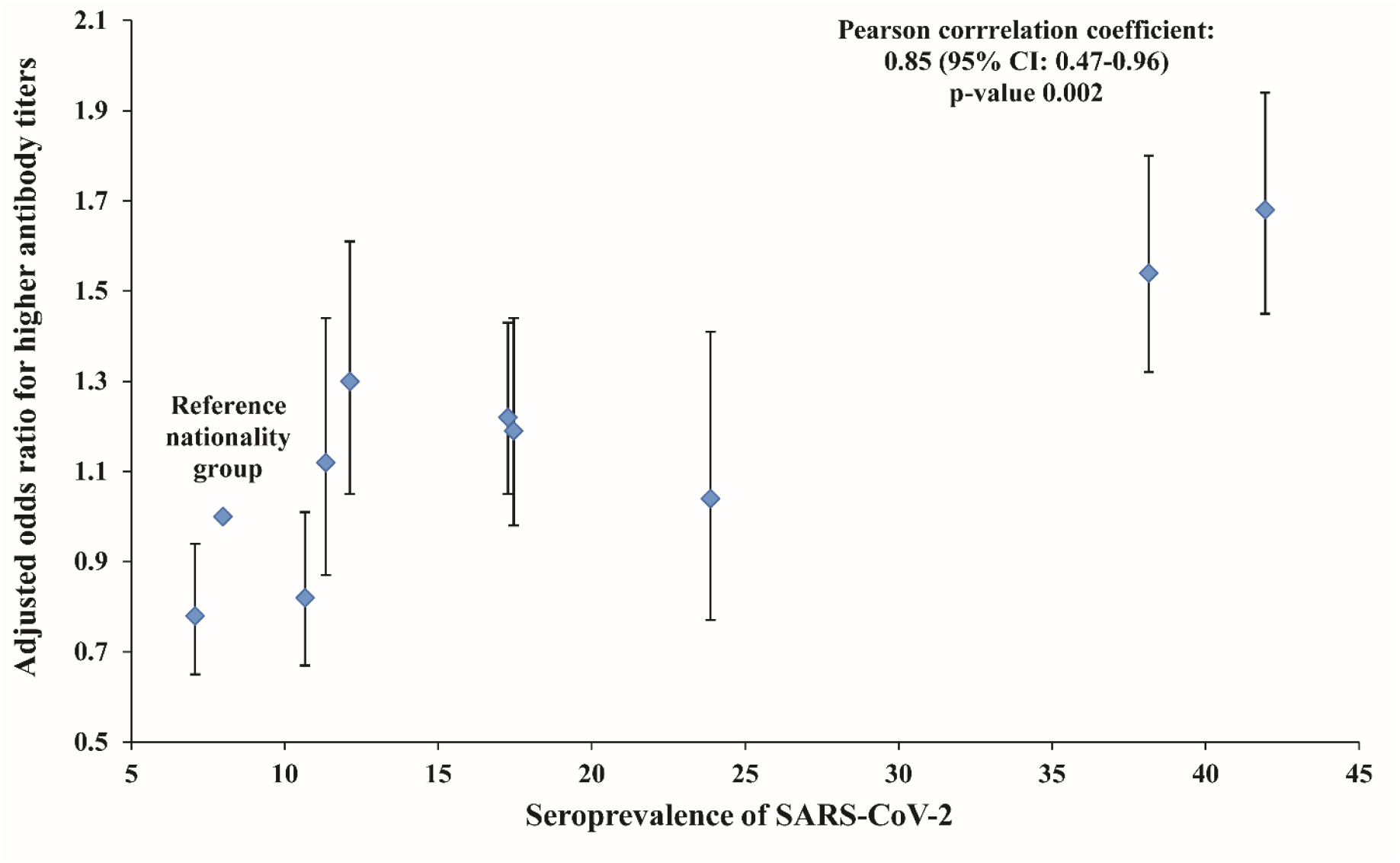
Adjusted odds ratios for higher antibody titers (optical densities higher than the median value of 27.0) for each nationality (Table 3) versus the corresponding SARS-CoV-2 seroprevalence for that nationality (Table 1).

Of the 18,844 antibody-positive persons, 9,375 had a PCR-confirmed diagnosis prior to the antibody-positive test—47.1% (95% CI: 46.1-48.2%) (Table 3). Meanwhile, 1,085 of the 18,844 antibody-positive persons had or progressed to a severe infection, and 393 had or progressed to critical infection. Thus, the infection severity rate was 3.9% (95% CI: 3.7-4.2%), the infection criticality rate was 1.3% (95% CI: 1.1-1.4%), and the combined infection severity or criticality rate was 5.2% (95% CI: 4.9-5.5%). With exactly 100 COVID-19 deaths recorded among the antibody-positive persons, the infection fatality rate was 0.3% (95% CI: 0.2-0.3%).

**Table 3.** Key SARS-CoV-2 epidemiological measures assessed in the study.

| **Epidemiological measure** | **Sample (denominator)** | **Positive for outcome (numerator)** | **Estimate in % (95% CI)** |
| --- | --- | --- | --- |
| Sample antibody positivity (seropositivity) | 112,941 | 18,844 | 16.7 (16.5-16.9) |
| Weighted antibody positivity (seropositivity) | 112,941 | 18,844 | 13.3 (13.1-13.6)* |
| Proportion with prior PCR-confirmed diagnosis<sup>†</sup> | 18,844 | 9,375 | 47.1 (46.1-48.2)* |
| Infection severity rate<sup>‡</sup> | 18,844 | 1,085 | 3.9 (3.7-4.2)* |
| Infection criticality rate<sup>§</sup> | 18,844 | 393 | 1.3 (1.1-1.4)* |
| Infection severity or criticality rate<sup>**</sup> | 18,844 | 1,478 | 5.2 (4.9-5.5)* |
| Infection fatality rate<sup>††</sup> | 18,844 | 100 | 0.3 (0.2-0.3)* |

PCR, polymerase chain reaction; CI, confidence interval.
\*Estimates weighted by sex, age, and nationality.
<sup>†</sup>Proportion of antibody-positive persons who had a prior SARS-CoV-2 PCR-confirmed diagnosis.
<sup>‡</sup>Number of infections that were severe per World Health Organization criteria [15] over the total number of antibody-positive persons.
<sup>§</sup>Number of infections that were critical per World Health Organization criteria [15] over the total number of antibody-positive persons.
<sup>\*\*</sup>Number of infections that were severe or critical per World Health Organization criteria [15] over the total number of antibody-positive persons.
<sup>††</sup>Number of COVID-19 deaths per World Health Organization criteria [15] over the total number of antibody-positive persons.

## Discussion

The above results indicate that <20% of the urban population of Qatar, which constitutes ∼40% of the total population and includes nearly all older adults, manifests evidence of prior infection, substantially less than the seroprevalence in the CMW part of the population, which was estimated recently in a nationwide survey at 55.3% [13].

This finding suggests that the lockdown and imposed social and physical distancing restrictions have been more successful in slowing transmission in the urban population compared to the CMW population. Building on the totality of evidence on the Qatar epidemic [11-13,20,21], this is possibly due to differences in the dwelling structure, in that the urban population lives mostly in single-unit or family households that each includes a small number of individuals. Meanwhile, the CMW population lives mostly in large shared accommodations that each includes a large number of individuals. While the lockdown forced individuals to stay more at home, it is typical to have more social contacts every day at home in the large shared accommodations than in the single-unit or family households, thereby reducing the options for effective social and physical distancing. This outcome highlights the role of the “boarding school” effect in respiratory infection transmission, seen often in the intense influenza outbreaks in regular and boarding schools [22,23]. This effect has been also seen in the large SARS-CoV-2 outbreaks in nursing homes in Europe and the United States [24-26].

With a seroprevalence of <20%, the urban population of Qatar remains far below the herd immunity threshold, estimated at 60-70% infection exposure [21,27,28]. Accordingly, there exists a potential for subsequent waves of infection in this part of the population, though no second wave has materialized since the epidemic peaked in late May, 2020, seven months ago [12,20]. On the contrary, only a slow increase in seroprevalence has occurred since the peak of the first wave (Table S1 of SI), reflecting the actual low incidence of infection in Qatar over the last few months [20]. The absence of a second wave, despite the lack of a lockdown and easing of many social distancing restrictions, may be explained by an “immunity shield” effect [29] arising from the social mixing between the urban and CMW populations, and by effective implementation of “*R*_*t*_ tuning”, an adjustment of restrictions based on the *R*_*t*_ value, by national policymakers, to prevent a second wave [20].

There were significant differences in seropositivity by sex, age, and nationality. These are probably not due to biological differences, but to differences in the likelihood of exposure to the infection. Indeed, a small proportion of the specimens tested in this study belonged to CMWs who had a higher risk of exposure to the infection than the urban population [13,21]. While HMC provides healthcare primarily to the urban population and other providers cater to the CMW population, HMC is the main tertiary care center in Qatar, and was also the nationally designated provider for COVID-19 healthcare needs. Thus, it is likely that a small proportion of specimens, which cannot be estimated precisely, was drawn from CMWs who were hospitalized for COVID-19 or other reasons. This may explain the higher antibody positivity of young Bangladeshi, Indian, and Nepalese men (Table 1), who form the bulk of the CMW population [13,21]. This may also explain the higher seroprevalence in the blood specimens drawn during inpatient or emergency clinical-care, which are more likely to be COVID-19-related, than those drawn during outpatient or home care/follow-up consultation clinical-care (Table 1). The higher exposure among men 20-69 years of age probably reflects their more frequent work and other activities outside the home, whereas men ≥70 years of age, urged through public-health messaging to remain at home, were more likely to do so, out of concern about infection severity.

The proportion of those antibody-positive who had a PCR-confirmed diagnosis prior to the antibody-positive test, was 47.1% (Table 3), much higher than the 9.3% in the CMW population [13], and that estimated for the total population of Qatar (11.6%) [20]. This is probably because study specimens were drawn from individuals receiving healthcare, including those hospitalized for COVID-19, people more likely to have been tested for the infection. This fact, along with the difference in age structure between the urban and CMW populations [4,8,9,13], may have resulted in higher estimates of infection severity, criticality, and fatality rates in this study (Table 3), compared to the study of the CMW population [13], or model predictions for the entire population of Qatar [30].

Strikingly, having a higher antibody titer varied by nationality, clinical-care type, and with time (Table 2). Variation by nationality is probably an indirect biomarker of re-exposure to infection, resulting in repeated immune-system reactivation. This is suggested by the very strong positive correlation between the odds of having a higher antibody titer and seroprevalence across the nationalities (Figure 2). Lower antibody titers were found in inpatients, but this may reflect COVID-19 hospitalizations for recent infections, so that there was not sufficient time for higher antibody titers to develop. There was a trend of increasing *higher antibody titers* over time, which may reflect the growing pool of infected persons who have had more time to develop higher levels of detectable antibodies post-infection, or alternatively to being re-exposed to the infection.

This study has some limitations. The sample included individuals attending HMC for routine or other clinical care, but this population may not necessarily be representative of the wider urban population of Qatar. Some specimens may have been drawn from CMWs, who are not representative of the urban population. However, the large sample size, equivalent to ∼10% of the urban population of Qatar, as well as the probabilistic weighting used in the analysis may have reduced inherent biases in our sample. Laboratory methods were based on high-quality, validated commercial platforms, such as the Roche platform used for serological testing [16,31]. The Roche platform is one of the most extensively used and investigated commercial platforms, with a specificity ≥99.8% [16,32,33] and a sensitivity ≥89% [12,31,33]. However, it is possible that the less-than-perfect sensitivity, especially for those with recent infections, may have underestimated the actual seroprevalence. Indeed, a recent investigation of the performance of three automated, commercial, serological platforms in Qatar, including the Roche platform, found that each of them missed ≥20% of individuals with past or current infections [34].

In conclusion, fewer than two in every 10 individuals in the urban population of Qatar had detectable antibodies against SARS-CoV-2, suggesting that this population is still well below the herd immunity threshold and is potentially at risk from a subsequent epidemic wave. This emphasizes the need to maintain current social and physical distancing restrictions while SARS-CoV-2 vaccinations are being scaled up throughout the country. The findings also suggest that higher antibody titers appear to be a biomarker of repeated exposures to the infection.

## Data Availability

All data are available within the manuscript and its supplementary materials.

## Acknowledgements

We thank Her Excellency Dr. Hanan Al Kuwari, Minister of Public Health, for her vision, guidance, leadership, and support. We also thank Dr. Saad Al Kaabi, Chair of the System Wide Incident Command and Control (SWICC) Committee for the COVID-19 national healthcare response, for his leadership, analytical insights, and for his instrumental role in enacting data information systems that made these studies possible. We further extend our appreciation to SWICC Committee and Scientific Reference and Research Taskforce (SRRT) members for their informative input, scientific technical advice, and enriching discussions. We also thank Dr. Mariam Abdulmalik, CEO of the Primary Health Care Corporation and the Chairperson of the Tactical Community Command Group on COVID-19, as well as members of this committee, for providing support to the teams that worked on the field surveillance. We further thank Dr. Nahla Afifi, Director of Qatar Biobank (QBB), Ms. Tasneem Al-Hamad, Ms. Eiman Al-Khayat and the rest of the QBB team for their unwavering support in retrieving and analyzing samples and in compiling and generating databases for COVID-19 infection, as well as Dr. Asmaa Al-Thani, Chairperson of the Qatar Genome Programme Committee and Board Vice Chairperson of QBB, for her leadership of this effort. We also acknowledge the dedicated efforts of the Clinical Coding Team and the COVID-19 Mortality Review Team, both at Hamad Medical Corporation, and the Surveillance Team at the Ministry of Public Health.

## Author contributions

PC conceived and designed this study and led its implementation and antibody testing. HC managed the databases, performed the statistical data analyses, and co-wrote the first draft of the manuscript. LJA led the statistical analyses and co-wrote the first draft of the article. All authors contributed to development of the study protocol, data collection, and acquisition, database development, discussions and interpretation of the results, and to the writing of the manuscript. All authors have read and approved the final manuscript.

## Supplementary Information

**Table S1.**
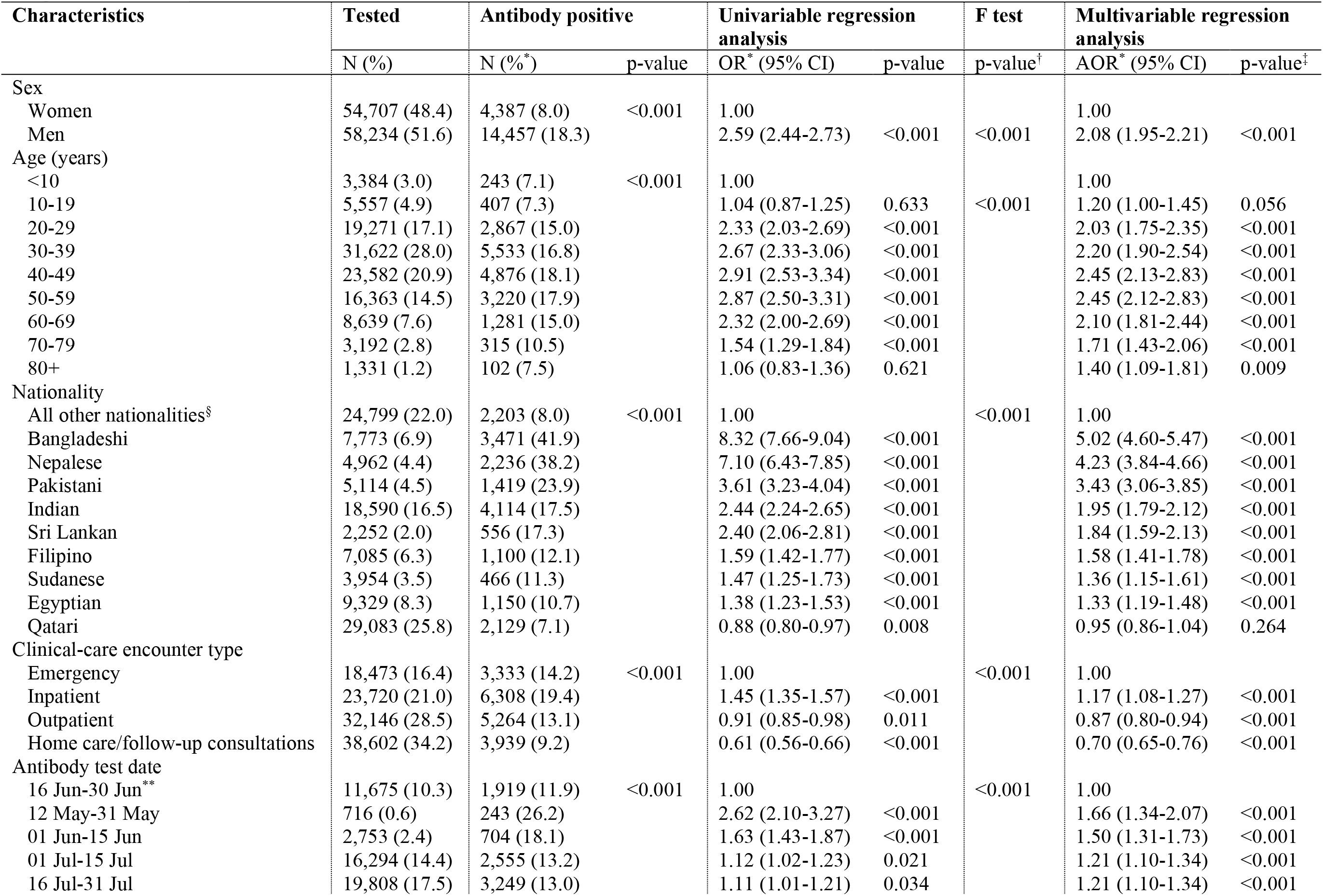
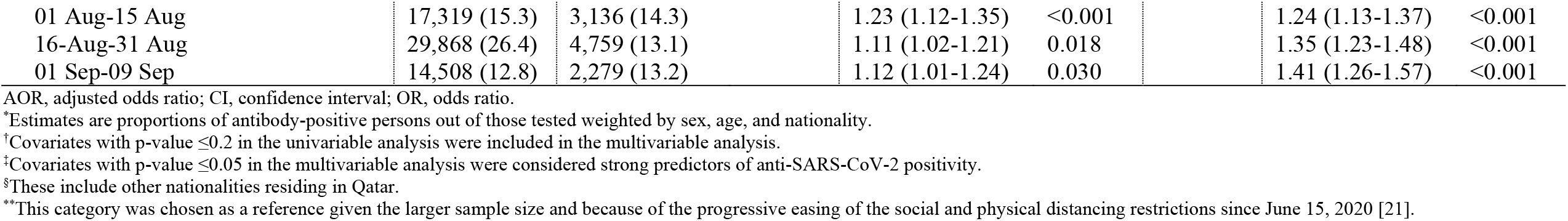
Results of regression analyses for the association with antibody positivity where antibody test date has been included as a categorical term.

**Table S2.**
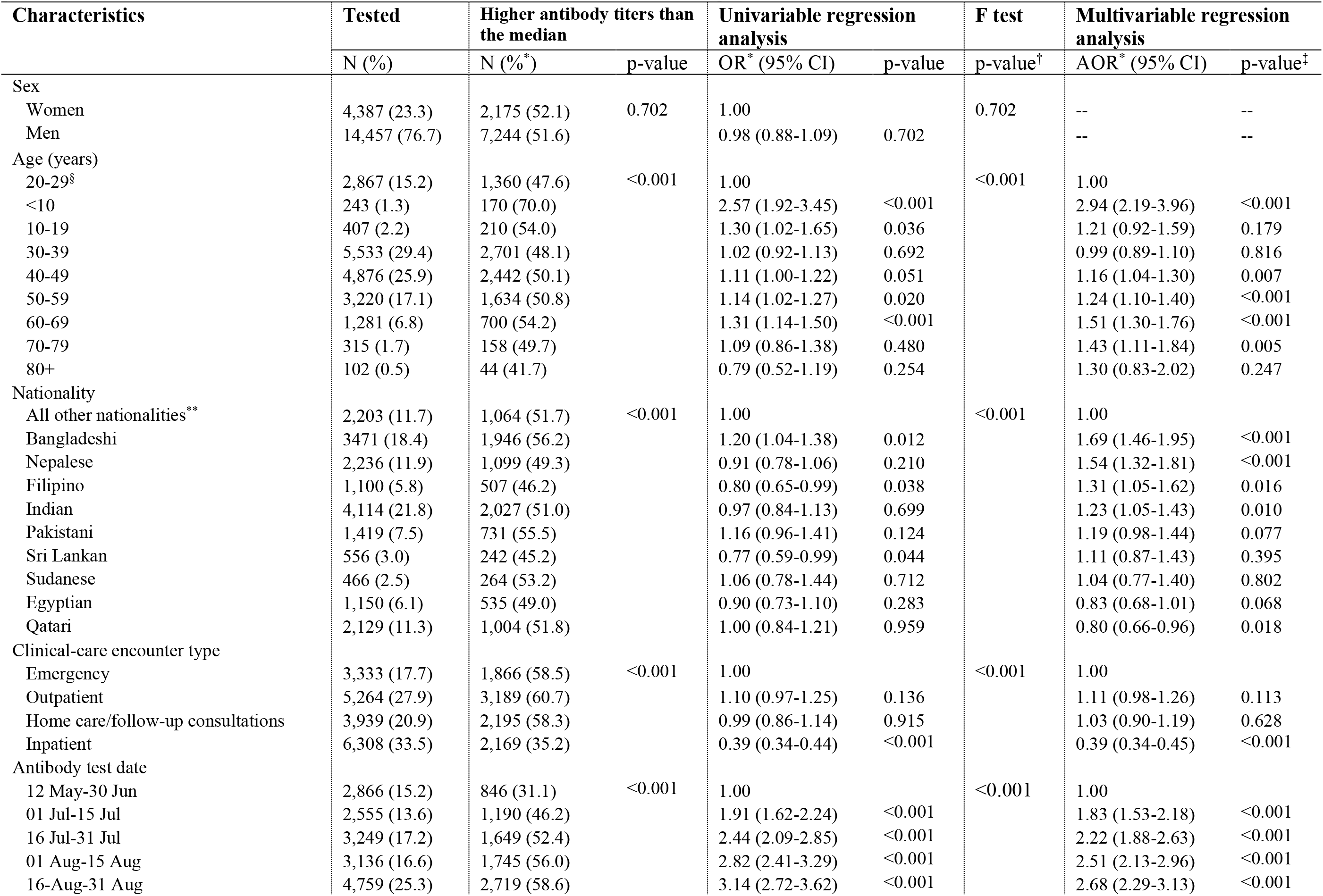
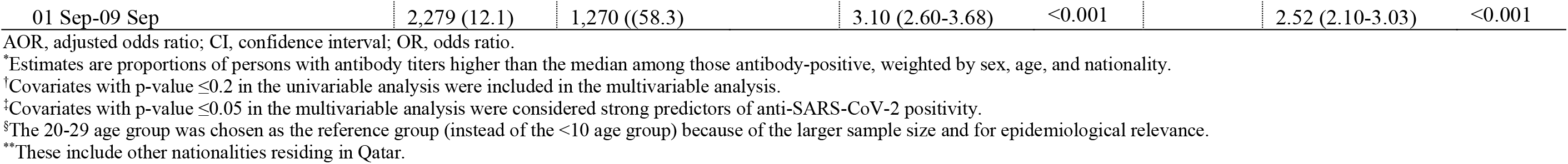
Associations for having a higher antibody titer (optical density) than the median value (≥27.0) among 18,844 antibody-positive individuals, where antibody test date has been included as a categorical term.

